# Natural language processing for patient selection in phase I/II oncology clinical trials

**DOI:** 10.1101/2021.02.07.21249271

**Authors:** Julie Delorme, Valentin Charvet, Muriel Wartelle, François Lion, Bruno Thuillier, Sandrine Mercier, Jean-Charles Soria, Mikael Azoulay, Benjamin Besse, Christophe Massard, Antoine Hollebecque, Loic Verlingue

**Affiliations:** Institut Optique Graduate School, Paris, France; Telecom Paris Tech, Paris, France; Department of Computing Science, University of Glasgow, Glasgow G12 8RZ; Informatic Team (DTNSI), Gustave Roussy, Villejuif, France; Drug Development Department (DITEP), Gustave Roussy, Villejuif, France; University Paris-Saclay, France; Medical Oncology Department, Gustave Roussy, Villejuif, France; INSERM UMR1030, Molecular Radiotherapy and Therapeutic Innovations, Gustave Roussy, Villejuif, France

## Abstract

**Purpose:** Early discontinuation affects over one-third of patients enrolled in early-phase oncology clinical trials. Early discontinuation is deleterious both for the patient and for the study, by inflating its duration and associated costs. We aimed at predicting the successful screening and dose-limiting toxicity period completion (SSD) from automatic analysis of consultation reports.

**Materials and methods:** We retrieved the consultation reports of patients included in phase I and/or phase II oncology trials for any tumor type at Gustave Roussy, France. We designed a pre-processing pipeline that transformed free-text into numerical vectors and gathered them into semantic clusters. These document-based semantic vectors were then fed into a machine learning model that we trained to output a binary prediction of SSD status.

**Results:** Between September, 2012 and July, 2020, 56,924 consultation reports were used to build the dictionary, and 1,858 phase I/II inclusion reports were used to train (75%), validate (15%) and test (15%) a Random Forest model. Pre-processing could efficiently cluster words with semantic proximity. On the unseen test cohort of 264 consultation reports, the performances of the model reached: F1 score 0.80, recall 0.81 and AUC 0.88. Using this model, we could have reduced the screen fail rate (including DLT period) from 39.8% to 12.8% (RR=0.322, 95%CI[0.209-0.498], p<0.0001) within the test cohort. Most important semantic clusters for predictions comprised words related to hematological malignancies, anatomo-pathological features and laboratory and imaging interpretation.

**Conclusion:** Machine learning with semantic conservation is a promising tool to assist physicians in selecting patients prone to achieve SSD in early-phase oncology clinical trials.

## Introduction

Phase I oncology trials aim at identifying the toxicity profile of first-in-human or newly developed anticancer drugs, hence selecting the correct dose for further investigations. Designs of the majority of phase I trials also include early assessment of clinical efficacy for multiple reasons, including (1) appropriate dose is not necessarily linearly correlated to toxicity ^1^, (2) early access drug approval programs, such as the PRIority MEdicines scheme (PRIME, EMA) and the Breakthrough Therapy designation (FDA), support optimization of early-phase trials and acceleration of clinical development, especially for drugs covering unmet medical needs, and (3) early assessment of drug efficacy can reduce development-associated costs ^2^. To ensure robust determination of tolerability and/or clinical efficacy upon early-phase trials, appropriate patient selection is critical. Patient selection is optimal when enrolled patients are more likely to benefit from the therapy and when they can be followed-up long enough to allow proper statistical analyses ^3,4^.

Patients entering phase I trials need to fulfill selection criteria, which are reviewed before the treatment starts, during a screening phase period that usually lasts one month (Figure 1). The rate of patients not eligible to the trial after this period – defining screening failure – usually scales between 10% and 30% ^5–7^. For patients who actually start the study treatment, there is an additional crucial period – defined as the dose-limiting toxicity (DLT) evaluation period – that usually prolongs of another 28 days the length of time during which if a patient is withdrawn from the trial and not evaluable for DLT (e.g. because of death or withdrawal of consent), he or she must be replaced. Thus, early discontinuation directly extends the duration of the study and majors development-associated costs. Altogether, these early discontinuation events have been described in approximately 40% of phase I patients and are mainly linked to detrimental cancer progression ^6^. Those are largely unpredictable by the current selection methods available.

**Figure 1.**
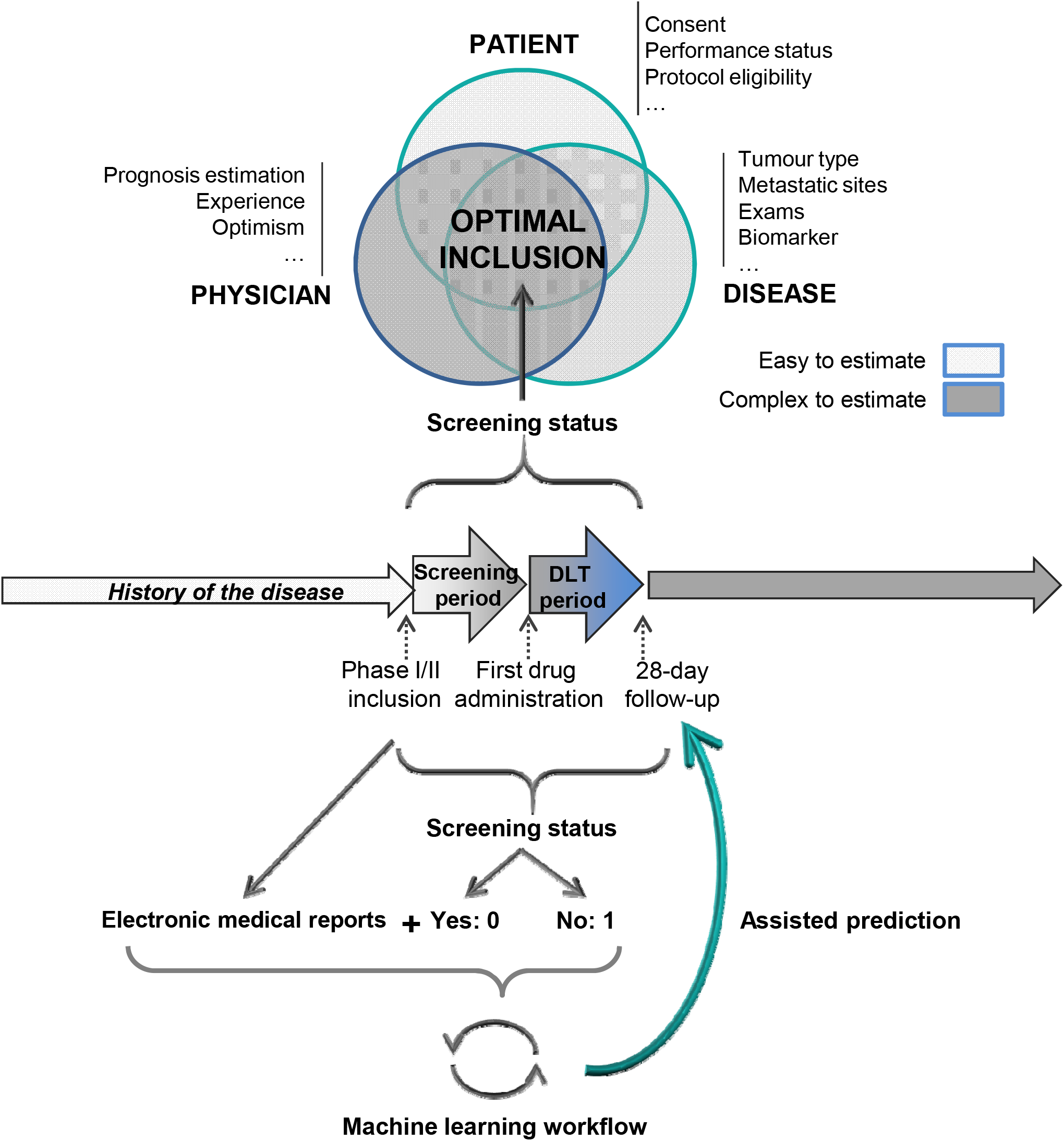
Abstract figure: human and machine learning could be complementary to estimate the successful screening and DLT period completion (screening status) of patients included in early phase clinical trials. At inclusion physician decision has to be quick and accurate, to provide the patient with a relevant proposition. Variables to estimate screening status are multiples, and are influenced by the physician, the patient (medical history composed of anti-tumor treatments and concomitant diseases) and the disease characteristics. The screening period usually lasts 3 weeks before the patient starts the investigational product. The Dose Limiting Toxicity (DLT) period usually lasts 28 days and is used to estimate the early tolerance of the investigational product and impacts further patients’ inclusion in the study. Usual causes of screen failures are rapid diseases progression, patient general status altered, concomitant disease(s) or new symptom(s), or new findings on screening exams that meet exclusion criteria. When tested on an independent cohort of 264 patients included in phase 1 oncology trial, our machine learning model could reduce the rate of screen failures from 39.8% to 12.8%.

Candidates for phase I clinical trials in oncology are usually patients affected by a metastatic or locally advanced cancer that previously progressed through all standard lines of treatments. Therefore, their prognosis, at the time of inclusion, is often complex to estimate. The rate of early non-drug related mortality during the first 90 days of phase I trials is estimated at around 16% of patients ^8^. This is of concern, not only because it affects the trial progress, but primarily because patients’ quality of end of life can be compromised by demanding trial procedures. To date, there is no accurate method to predict the early mortality risk and spare those patients. Clinicians have developed several tools that are regularly used to estimate patient survival – such as the Royal Marsden Hospital (RMH) or the Gustave Roussy immune (GRIm) scores – although high rates of early-trial discontinuation persist in routine ^9,10^. Notably, a high RMH score (poor prognosis) was not significantly related to a higher risk of screen failure within a cohort of 1,293 phase I patients (OR = 2.3; 95% CI [1.0-5.7], p = 0.06) ^6^. Another study revealed that patient selection on RMH score reduced non-drug related 90-day mortality by 50%, but more than half of discarded patients would in fact have survived beyond 90 days, which highlights the low specificity of this method ^8^.

The deployment of electronic health records has recently driven many applications using machine and deep learning ^11^. We therefore hypothesized that a machine learning model may be able to predict the outcome of the screening period based on free-text medical reports, and thereby help physicians to limit early trial discontinuation. Using a large base of consultation reports from Gustave Roussy’s electronic health records, we developed and benchmarked a machine learning model that aimed to predict successful screening and DLT period completion (SSD) for patients included in early-phase oncology clinical trials.

## Materials and Methods

### Selection criteria

Electronic health records selected for this study referred to all the patients enrolled between September, 2012 and July, 2020 in a phase I (including first-in-human) or phase II for any tumor type, at the Drug Development Department (DITEP) or at the Medical Oncology Department (DMO) for lung cancer patients, at Gustave Roussy, France. These selection criteria were supported by internal authorizations and the deployment of systematic electronic recording at DITEP.

### Corpus

Text documents were collected retrospectively from the electronic report data center at Gustave Roussy. The global text corpus (TC1) comprised all the long reports related to the selected patients. Long reports were defined as reports comprising at least 250 characters. The restricted text corpus (TC2) comprised only consultation reports dedicated to inclusion in a clinical trial (one record per inclusion per patient).

Consultation reports were semi-structured documents containing free-text in French. Contents were classical medical observations usually organized following medical logic, such as: description of the reason of consultation or hospitalization, antecedents, risk factors and lifestyle, history of the disease, daily treatments, physical examination, interpretation of other exams and a conclusion, among other non-systematic information such as description of the therapeutic procedures.

We used a random split rule on the TC2 to isolate 3 independent cohorts: 70% for the training cohort, 15% for the validation cohort and 15% for the test cohort. The validation cohort was used to train the learning hyper-parameters and to benchmark the model’s performances. The test cohort was used to independently evaluate the generalization performance of the predictions. Reports used for the test cohort were removed from TC1 to limit potential data leakage.

### Free-text processing

We designed a natural language pre-processing pipeline to turn French free-text into numerical features that could be used to feed a Random Forest model. The working dictionary was composed by the all words retrieved from TC1. This dictionary was lemmatized, tokenized, vectorised, and reduced to semantic clusters. Lemmatization corresponds to the reduction of words to their root form ^12 13^. With lemmatization, inflected words remained compatible with language. To perform lemmatization on TC1, we used a convolutional network pre-trained on the UD French Sequoia v2.5 and WikiNER datasets from the python spaCy library. After tokenization, words were reduced to a vector of indexed objects called *tokens* ^14^. For text cleaning, we removed irrelevant elements such as *html* marks and French stop-words (corresponding to *the, in, a, at, and, of*, etc.). Cleaning of numerical values remains controversial and lacks clear guidelines to our knowledge. We considered that the list of numerical values taken as characters was disproportionately large. Moreover, while some numbers can be useful – such as clinical information on the patient (weight, age etc.) and treatments (doses, schedules, dates, etc.) – others are major sources of noise, such as some dates or patient’s phone number. We thus decided to remove numerical data.

### Word embedding

Word embedding consists in representing words (character lists) with numerical vectors to use it as inputs for machine learning models. Word2Vec has the advantage of conserving words’ semantic meaning. Word2Vec is a two-layers neural network that we trained using the “skip-gram” option (from a word, the model’s task was to predict the context, i.e. the n-grams surrounding words) ^15^. The hidden layer size was arbitrarily set to 120 and used to generate numerical embedding vectors.

### Dimensionality reduction

Dimensionality reduction consists in reducing the number of features within the data while conserving some of their properties. We used UMAP (Uniform Manifold Approximation and Projection). UMAP preserves point-wise distances from high-dimensional space to a low-dimensional one ^16^.

### Clustering

We performed clustering to extract the information generated by UMAP dimension reduction. We used Agglomerative Clustering, a non-supervised hierarchical clustering method to build nested clusters by merging or dividing them successively ^17^. This approach minimizes the variance upon clusters. The number of clusters is user-defined.

### Vectors of cluster frequencies

The clusters were used to build one vector per report whose length corresponded to the number of clusters and values to the normalized frequency of words that belong to each cluster. To do so, each word detected within a report was replaced by the index of its related cluster. Frequencies of cluster indexes were normalized by the total sum of the words in the corresponding report. After ranking by cluster indexes, the vector was fed to the machine learning model.

### Modeling

#### Learning model

Random Forest uses an ensemble of decision trees for prediction ^18^. The input features constituted the nodes of the trees and cutoff values were randomly defined. The final class was chosen by averaging the predictions of each tree in the forest. Running multiple forests with bootstrapping was used to select the best forest.

#### Labels

The task was to predict a binary label: class 0 in case of successful screening and DLT period completion, class 1 in case of screen failure. Successful screening and DLT period completion concerned patients who remained included in the trial 28 days after the first drug administration. Consequently, screen failures concerned patients who were withdrawn from study during either the screening period (between inclusion and start of treatment) or the DLT period.

#### Performance metrics

We estimated the performances of the model by calculating the Area Under the Receiver Operating Characteristic (ROC)Curve (AUC), the recall and F1 scores. The ROC curve was generated by computing the true positive rates (y-axis) *versus* the false positive rates (x-axis) for multiple cutoff values ^19^. Recall (also called sensitivity) was the ratio of the number of reports correctly assigned to class 1 (true positives) divided by the total number of reports truly belonging to class 1 (true positives plus false negatives). Precision (or accuracy) was the ratio of true positives among true positives plus false positives. The F1 score was calculated by the harmonic mean of precision and recall, with formula: F1 = 2X(precisionXrecall)/(precision+recall). For each score, the maximum value is 1 and corresponds to perfect prediction. Scores > 0.5 are better than random, and we considered scores > 0.8 as highly predictive.

To further compare the predicted screen failure rate obtained from the algorithm with the observed screen failure rate within the final test cohort, we used the Pearson’s chi-squared test. The relative risk (RR), its standard error and 95% confidence interval (95%CI) were calculated according to *Altman DG, 1991*. We considered p<0.05 to be statistically relevant.

### Cross-validation and test

We used cross-validation procedure to select the best hyper-parameters of the pre-processing pipeline and machine learning model. Hyper-parameters trained included: the number of clusters, number of trees in the forest, the number of features to consider when looking for the best split and the minimum number of samples required to be at a leaf node. We performed grid search on Random Forest F1 scores from the validation set to select the best final model hyper-parameters. We evaluated the generalization performance of the best validated model on the test cohort from TC2, as described above.

### Interpretation

To interpret the predictions of the random forest model, we used SHapley Additive exPlanations (SHAP) values that quantify the impact of each feature on the model outputs ^20^. To do so, all permutations in input features were generated. As thousands of trees are built in a random forest, various permutations of input features were available. These combinations were used to estimate the marginal contribution of each feature, and their weighted sum gave the corresponding SHAP value for a feature.

### Computational tools

The full code and notebook are accessible at https://github.com/DITEP/NLP_for_ScreenFail_prediction. We used JupyterLab environment for development with Python 3.6.10. For basic computations, we used Pandas 1.0.3 and Numpy 1.18.2. For most machine learning methods such as grid search, cross-validation and clustering, we used scikit-learn 0.22.2 ^21^. We used SpaCy 2.2.4, an open source software library for lemmatization in French. For tokenisation and lemmatization we loaded the language object fr_core_news_sm ^22^. We used Gensim library for word embedding with Word2Vec. The two libraries used for data visualization were Matplotlib 3.2.1 and Seaborn 0.10.0.

### Ethics

Patients included in this study were informed of the analysis of their personal data. The study was compliant with the General Data Protection and the French Law. The data collected could only be used for the aim of this study, and patient identification was secured. The study was conducted according to Good Clinical Practice Guidelines of the International Conference on Harmonization and the Declaration of Helsinki recommendations.

## Results

### Data

We retrospectively retrieved all electronic records of patients included in phase I/II oncology trials for any type of tumor at DITEP and for lung cancers at DMO. We considered reports recorded between September, 2012 and July, 2020, at Gustave Roussy, France. Among 111,091 documents, 56,924 were long reports (TC1), including 1,858 records that related to consultations dedicated to inclusions in early-phase clinical trials (TC2) (Figure 2). To allow the complete independence of the final test dataset, we excluded those reports from TC1. Patients entering early-phase clinical trials were affected by cancers of various types, either solid or hematological, usually at the refractory stage. Previous treatments received were heterogeneous, mostly chemotherapies, targeted therapies and immunotherapies.

**Figure 2.**
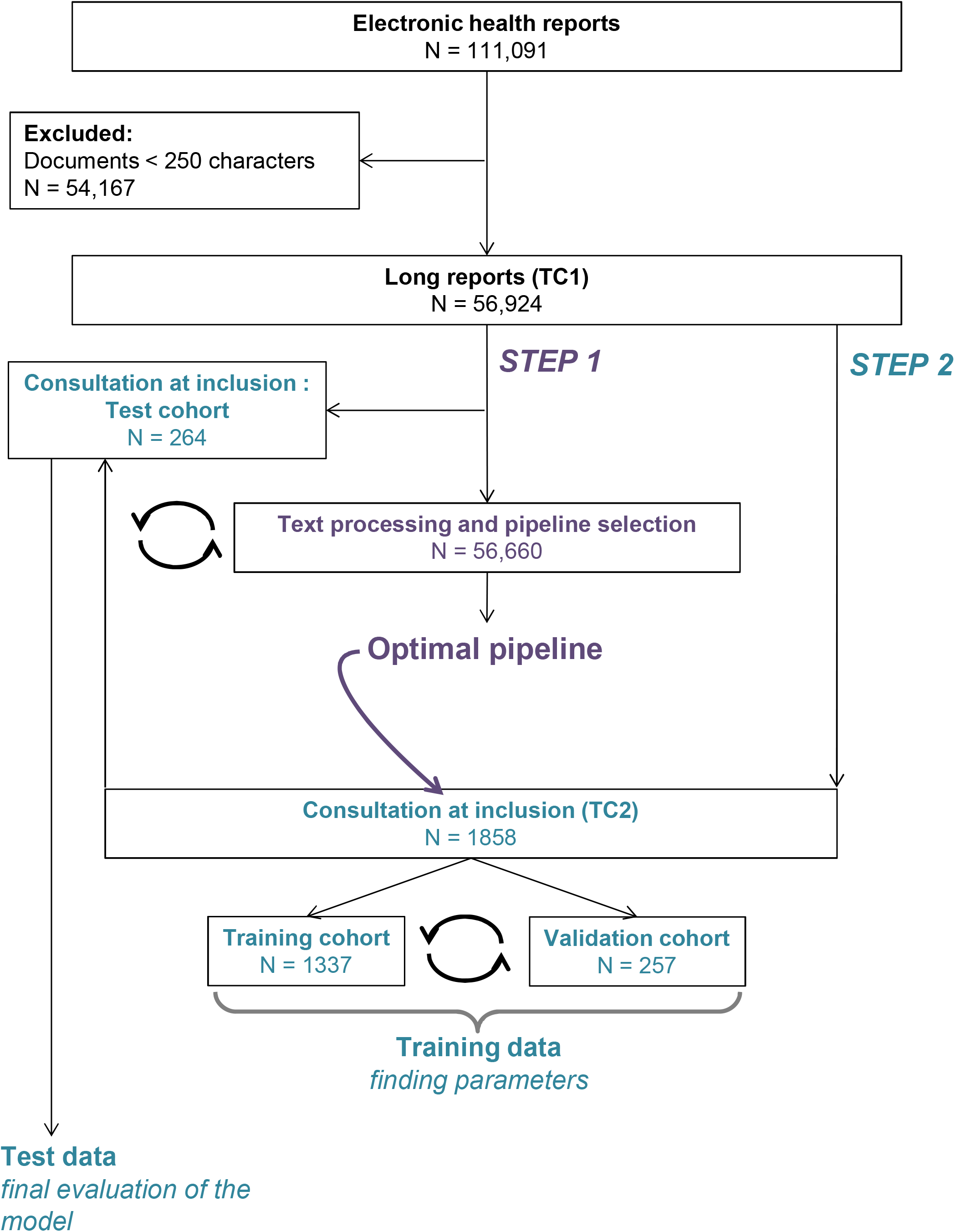
Flow chart and data split. TC1: text corpus 1, composed of all reports containing more than 250 characters (defining “long” reports); TC2: text corpus 2, composed of reports of the consultations dedicated to inclusion into phase I trials. TC2 was used to train, validate and test the machine learning model.

### From embedded words to clustered data

TC1 was used to optimize the pre-processing and to build the dictionary, including by word embedding with Word2Vec (Figure 3). We then reduced the dimensions of the word-embedded dictionary using UMAP with the “cosine” metrics (Figure 4A). On the resulting 2-dimensions matrix, we performed a supervised clustering to define 200 clusters (Figure 4B). At this stage, we could observe that combining such embedding and clustering methods was sufficient to identify groups of words with semantic similarities, such as words related to biopsies, clinical trials or radiography (figure 4C).

**Figure 3.**
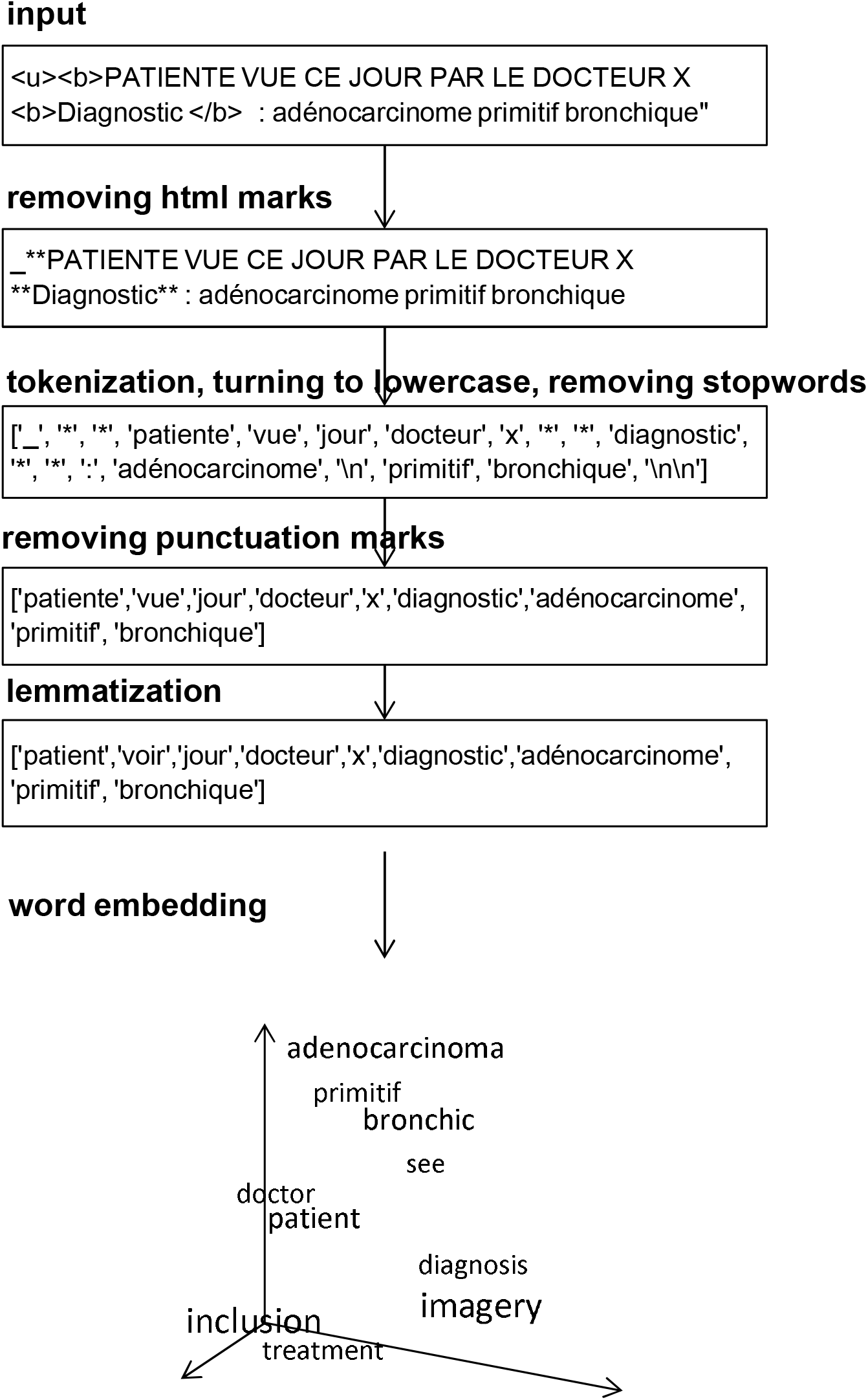
Example of free-text preprocessing used in this study and vectorization of its components into numerical data.

**Figure 4.**
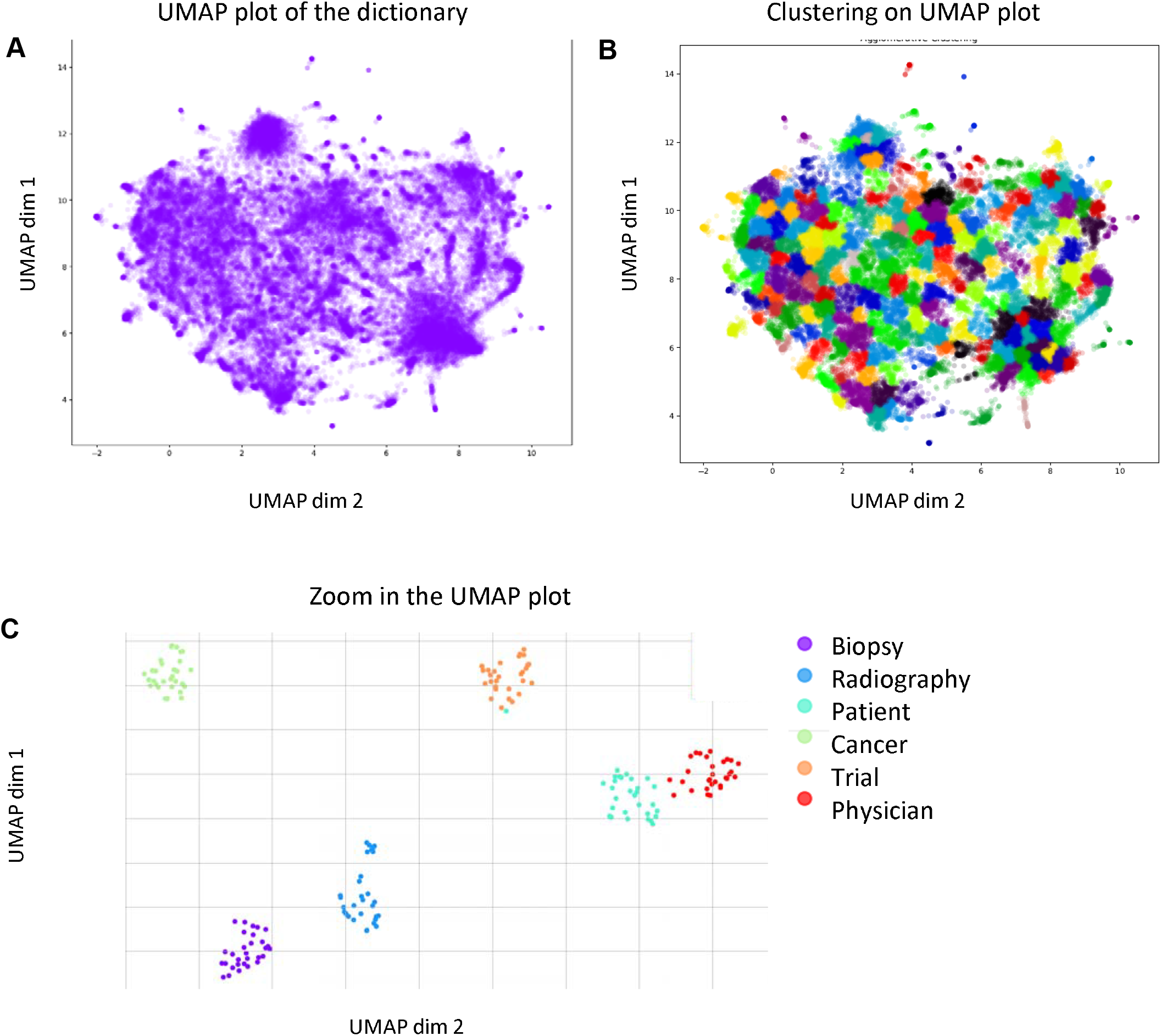
Clustering of the dictionary after dimensionality reduction of the word vectors. (**A**) UMAP 2D projection of TC1 embedding dictionary built from 56,638 documents. At least two levels of granularity appeared in the figure: a fine granularity with clusters of few words and a coarse granularity with clusters of many more words. (**B**) Supervised clustering of the UMAP matrix with 200 clusters. Clustering has been performed using the agglomerative clustering method. Most clusters, including peripheral ones that are easy to visualize, were well delineated. (**C**) Zoom in the UMAP clustering on the 30 most similar words associated with 6 items arbitrarily selected. We used the most_similar function in Word2Vec. Clustering was efficient even in the center of the UMAP cloud.

We thus transformed all free-text records from TC2 into 200-dimensions vectors where each vector value relates the frequency of the clusters within each report. For example, if one cluster comprised the words “disease”, “cancer”, “adenocarcinoma” and “NCSLC”, the vector value for the corresponding cluster was the number of times those words appeared in one report divided by the total number of words in the report.

### Results of the modeling

We trained a Random Forest model to predict the SSD from consultation reports at inclusion, using vectors of semantic clusters’ frequencies. We used a 5-fold cross-validation procedure on the training and validation sets to estimate intermediate generalization performance for hyper-parameter search (Figure 5A). Best hyper-parameters from the cross-validation procedure were obtained with 187 trees in the forest, 40 features per tree, one as the minimum number of samples required to be at a leaf node and considering from 100 to 200 distinct clusters for preprocessing (figure 5B). The best model reached the following validation performances: F1 score 1.00, recall 0.99 and AUC score 1.00.

**Figure 5.**
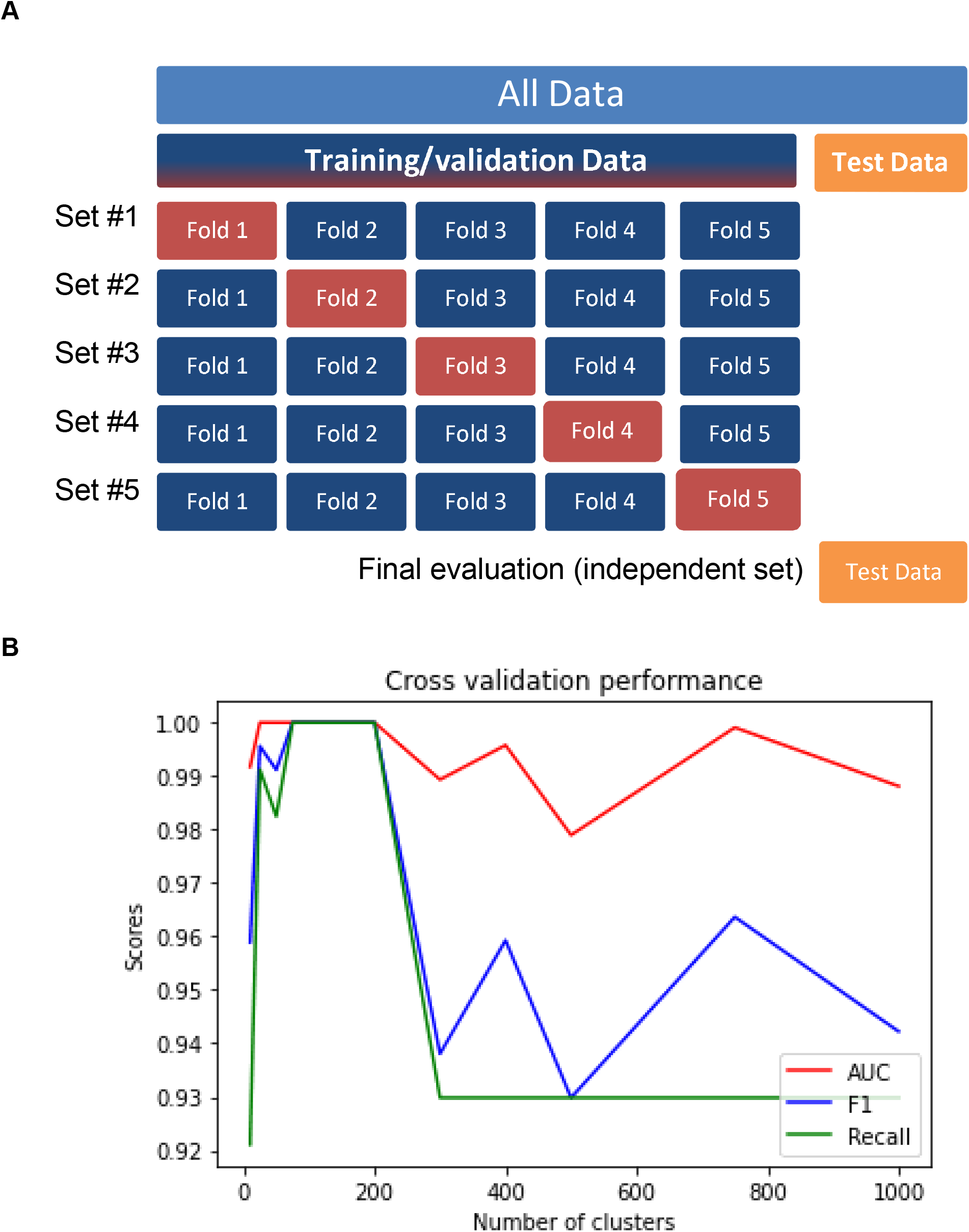
Cross-validation of the random forest model. (**A**) Illustration of the cross validation procedure. (**B**) Cross-validation performance of the random forest model given the number of clusters selected in the pre-processing. This observation supported our choice of 200 clusters for further analysis. AUC: Area Under the Receiver Operating Characteristic Curve

### Testing on unseen dataset

By testing our model on an independent and unseen dataset of 264 phase I consultation reports, the performances of the model reached: F1 score 0.80, recall 0.81 and AUC score 0.88 (Figure 6A). The confusion matrix showed that among 264 consultation reports, 136 patients with successful screening and DLT period completion were accurately predicted by the model (negative predictive value = 0.87), and 85 patients who experienced screen failure were also accurately predicted by the model (positive predictive value = 0.79) (Figure 6B). In our test cohort, using such a model-based prediction would have therefore reduced the rate of screen failure from 39.8% to 12.8% (RR=0.322, 95%CI [0.209-0.498], p<0.0001) (Figure 6C).

**Figure 6.**
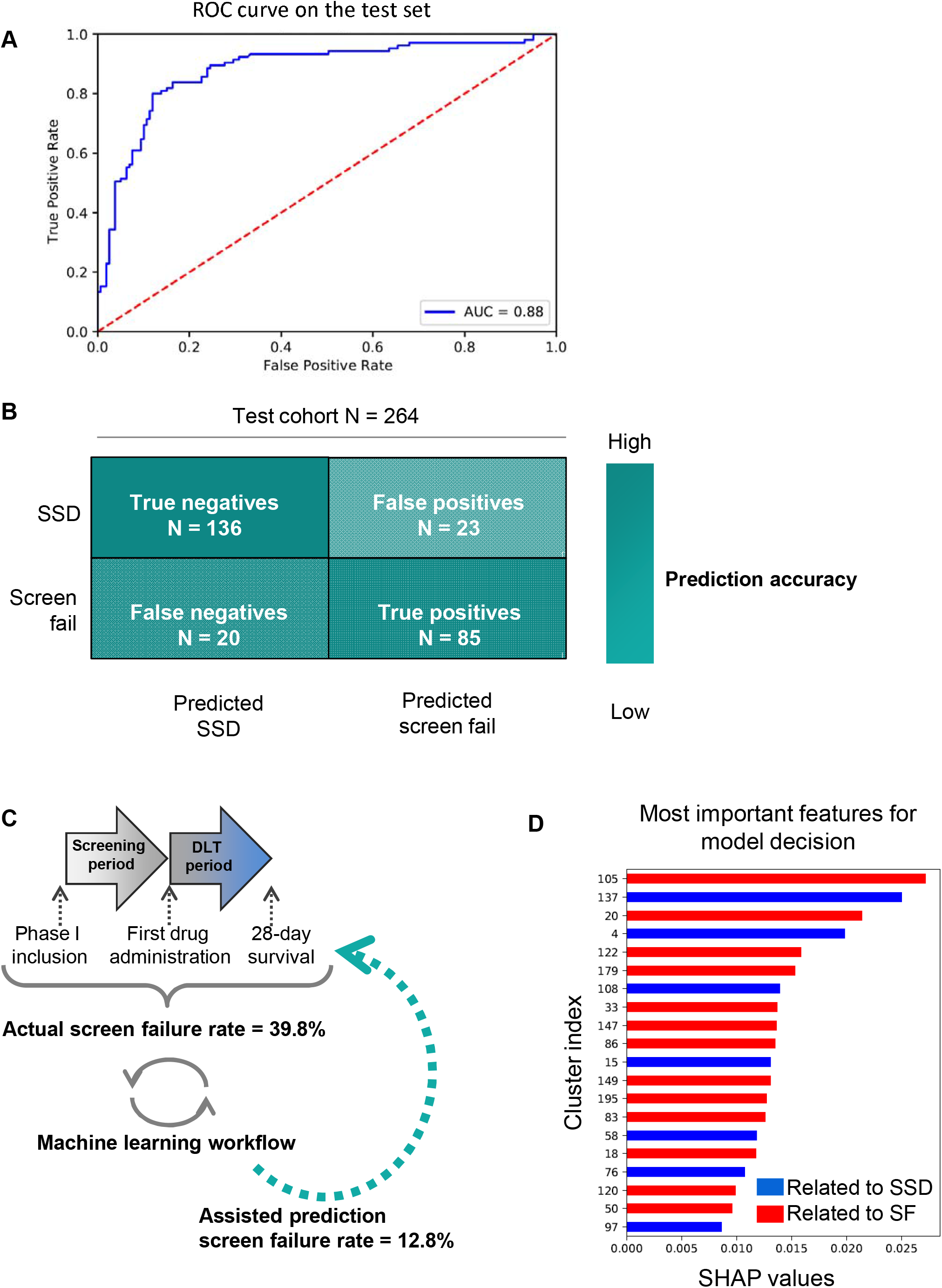
Random forest predictions on the independent and unseen test cohort. (**A**) Receiver Operating Characteristic (ROC) Curve. (**B)** Confusion matrix. (**C**) Comparison of human-based and machine-based screening success prediction. (D) Most important features for the model decision using SHAP values. SSD: successful screening and DLT period completion; SF: screen failure. AUC: Area Under the Curve, SSD: successful screening and DLT period completion

### Interpretation

We used the SHAP values to estimate the contribution of each input feature to the output of the model. The three most important semantic features were represented by clusters #105, #137 and #20 (Figure 6D). Words in cluster #105 referred to hematological malignancies, cluster 137 to anatomo-pathological specialties and cluster 20 to laboratory and imaging exams and results, including abnormal ones. Cluster #137 had a positive impact on the probability of SSD (the more frequently the words of those clusters appeared in the report, the more likely the patient was predicted to be a good candidate for inclusion). By contrast, cluster #105 and #20 were associated with a higher probability of screen failure prediction. To confirm our findings, we compared the frequency of each cluster in reports of patients associated with screen failure *versus* their frequency in reports of patients associated with SSD. We found that, amongst the 200 predefined clusters, 105 (52.5%) were significantly different according the two label classes (t-test p-value < 0.05).

## Discussion

We developed a pre-processing pipeline that converted free-text medical data to numerical vectors compatible with a machine learning model that predicted efficiently the initial outcome of patients included in early-phase clinical trials. Our Random Forest model achieved significant overall performances, with an AUC of 0.88 on the test cohort, and have reduced by three the rate of screen failure from 39.8% to 12.8% in a cohort composed of patients included in pan-tumor phase I/II clinical trials and lung cancer phase II clinical trials in our center.

Early-phase clinical trials are a cornerstone of larger improvement of the whole drug development process ^23–25^. Our previous experience showed that machine learning tools could predict the likelihood of FDA approval based on data extracted from phase I related published PubMed abstracts, combined with pharmacological information ^26^. Similarly, proper patient selection in early-phase clinical trials should improve the overall clinical drug development quality ^23,24^.

Selection criteria of early-phase clinical trials usually rely on biological values and performance status. We could not directly compare survival scores (RMH or GRIm) to our model prediction which is a binary screening status, independent from the time. However, in a cohort similar to ours, RMH score wrongly discarded half of potential good candidates for inclusion in early-phase clinical trials ^6^. This is of major concern, because early-phase clinical trials was shown to improve clinical outcomes for up to 30% of patients ^27^. Conversely, some patients may have benefited from alternative treatments or adapted supportive care if advocated earlier, with improved quality of life, reduced end-of-life complexity and perhaps improved survival. In our cohort, the rate of false positives, which refers to the fraction of patients that would have been wrongly predicted by the model as screen failure while they would have completed the screening and DLT period, was of 8.7%, which largely outperforms RMH score calculation.

Consultations at inclusion usually gather the most complete and most recent clinical observations summarizing patient’s history, recent clinical events and recent exams. This type of data is obviously knowledge-driven and the closest to screening status in one patient’s timeframe. This is also the most relevant time point as it is when patients provide their consent to participate to the clinical trial. The physician decision at this consultation has to be quick and accurate, to offer the patient a relevant treatment proposition while minimizing the risk of false expectations, which is an everyday challenge.

The screen failure rate observed in this study (including screening and DLT period) may seem high (about 40%), but was consistent with previous reports and can be explained by the known tendency of physicians to be overoptimistic regarding their patients prognosis ^7,28^. In this situation, using a machine learning tool to support the decision has the advantages (i) to be prompt, (ii) to take into account a large number of variables and (iii) to alleviate the weight and mental impact of the decision.

Although the performances of our model are satisfying, this study has limitations. First, records were retrospectively collected in a single center, and even if the reports were authored by multiple investigators, it is not certified that the model would accurately generalize to other hospital records. Patients in phase II clinical trials for lung cancer contributed to enlarge the scope of the training data. Other specific tumor types or later stage clinical trials could help complete the current tool. Also, it is restricted to French language. Second, this tool uses free text reports only, and does not support structured data format to date (e.g. biological values, performance status, ESCAT classification for efficacy prediction for targeted treatments, among others interesting measurements).

Third, our strategy neither takes into account the order of the words within the sentence (for example, the negations), known as semantic dependencies, nor the document structure. Recurrent neural networks can be used to consider semantic dependencies, but transformers that recently achieved the best performances in most natural language processing tasks do not clearly have this specificity ^29,30^. Finally, we chose a relatively shallow machine learning model: compared to deep learning models, Random Forest has less parameters to train. This was an advantage for our task with few numbers of examples and a large number of dimensions (number of words in the dictionary). This notably allowed decent generalization performance while deep learning models would have certainly overfitted the training cohort. Larger text corpus related to our task could be used to limit overfitting.

In conclusion, we developed a natural language processing pipeline that successfully produced semantic-derived vectors used to feed a machine learning model with accurate prediction of patients’ outcome during screening and DLT period in early-phase oncology clinical trials. This could be a useful tool to support decision of patient inclusion proposal and reduce the risk of screen failure and early discontinuation. We actively work on a prospective non-interventional validation of this study.

## Data Availability

Our data is based on personal medical information with risk of identification. We are not allowed to publically disclose it.

https://github.com/DITEP/NLP_for_ScreenFail_prediction

## Acknowledgments

The authors thank Daphné Morel (PharmD, PhD), for providing medical writing and editorial support. The authors also acknowledge the DTNSI team, including Yannick Boursin, Emmanuel Planchet, Olfa Makkaoui, the DITEP Data Science team, including Brice Aouchiche, Guillaume Beinse, Virgile Tellier, Ugo Benassayag, Hichem Alaoui, Elise Nassif, Hélène Vanacker, Frédéric Bigot, clinicians and researchers at Gustave Roussy, including Samy Amari, Eric Angevin, Fabrice André, Stefan Michiels, Eric Deutsch, researchers at CentraleSupelect, including Sarah Lemler, Paul-Henry Cournède, Veronique Letort and Enrico Sartor.

## Conflict of interest

LV reports personal fees from Adaptherapy, grants from Bristol-Myers Squibb, non-personal fees from Servier and Pierre-Fabre, outside of the submitted work.

LV, AH & CM, as part of the Drug Development Department (DITEP):

Principal/sub-Investigator of Clinical Trials for Abbvie, Adaptimmune, Aduro Biotech, Agios Pharmaceuticals, Amgen, Argen-X Bvba, Arno Therapeutics, Astex Pharmaceuticals, Astra Zeneca Ab, Aveo, Basilea Pharmaceutica International Ltd, Bayer Healthcare Ag, Bbb Technologies Bv, Beigene, Blueprint Medicines, Boehringer Ingelheim, Boston Pharmaceuticals, Bristol Myers Squibb, Ca, Celgene Corporation, Chugai Pharmaceutical Co, Clovis Oncology, Cullinan-Apollo, Daiichi Sankyo, Debiopharm, Eisai, Eisai Limited, Eli Lilly, Exelixis, Faron Pharmaceuticals Ltd, Forma Tharapeutics, Gamamabs, Genentech, Glaxosmithkline, H3 Biomedicine, Hoffmann La Roche Ag, Imcheck Therapeutics, Innate Pharma, Institut De Recherche Pierre Fabre, Iris Servier, Janssen Cilag, Janssen Research Foundation, Kura Oncology, Kyowa Kirin Pharm. Dev, Lilly France, Loxo Oncology, Lytix Biopharma As, Medimmune, Menarini Ricerche, Merck Sharp & Dohme Chibret, Merrimack Pharmaceuticals, Merus, Millennium Pharmaceuticals, Molecular Partners Ag, Nanobiotix, Nektar Therapeutics, Novartis Pharma, Octimet Oncology Nv, Oncoethix, Oncopeptides, Orion Pharma, Ose Pharma, Pfizer, Pharma Mar, Pierre Fabre, Medicament, Roche, Sanofi Aventis, Seattle Genetics, Sotio A.S, Syros Pharmaceuticals, Taiho Pharma, Tesaro, Xencor. Research Grants from Astrazeneca, BMS, Boehringer Ingelheim, Janssen Cilag, Merck, Novartis, Onxeo, Pfizer, Roche, Sanofi. Non-financial support (drug supplied) from Astrazeneca, Bayer, BMS, Boringher Ingelheim, Medimmune, Merck, NH TherAGuiX, Onxeo, Pfizer, Roche.

The other authors have no conflict of interest to declare.

## References

1. Postel-Vinay, S. et al. Challenges of phase 1 clinical trials evaluating immune checkpoint-targeted antibodies. Ann. Oncol. 27, 214–224 (2016).

2. Nass, S. J. et al. Accelerating anticancer drug development - opportunities and trade-offs. Nat Rev Clin Oncol 15, 777–786 (2018).

3. Chakiba, C., Grellety, T., Bellera, C. & Italiano, A. Encouraging Trends in Modern Phase 1 Oncology Trials. New England Journal of Medicine 378, 2242–2243 (2018).

4. Schwaederle, M. et al. Association of Biomarker-Based Treatment Strategies With Response Rates and Progression-Free Survival in Refractory Malignant Neoplasms: A Meta-analysis. JAMA Oncol (2016) doi:10.1001/jamaoncol.2016.2129.

5. Mckane, A. et al. Determinants of patient screen failures in Phase 1 clinical trials. Invest New Drugs 31, 774–779 (2013).

6. Kempf, E. et al. A Case-Control Study Brings to Light the Causes of Screen Failures in Phase 1 Cancer Clinical Trials. PLoS ONE 11, e0154895 (2016).

7. Fogel, D. B. Factors associated with clinical trials that fail and opportunities for improving the likelihood of success: A review. Contemp Clin Trials Commun 11, 156–164 (2018).

8. Olmos, D. et al. Patient selection for oncology phase I trials: a multi-institutional study of prognostic factors. J. Clin. Oncol. 30, 996–1004 (2012).

9. Arkenau, H.-T. et al. Clinical outcome and prognostic factors for patients treated within the context of a phase I study: the Royal Marsden Hospital experience. Br. J. Cancer 98, 1029–1033 (2008).

10. Bigot, F. et al. Prospective validation of a prognostic score for patients in immunotherapy phase I trials: The Gustave Roussy Immune Score (GRIm-Score). Eur. J. Cancer 84, 212–218 (2017).

11. Shickel, B., Tighe, P., Bihorac, A. & Rashidi, P. Deep EHR: A Survey of Recent Advances in Deep Learning Techniques for Electronic Health Record (EHR) Analysis. IEEE Journal of Biomedical and Health Informatics 22, 1589–1604 (2018).

12. Labbé, D. La lemmatisation des grandes bases de textes. Un exemplel: Corneille, Molière et Racine. (2002).

13. Stemming and lemmatization. https://nlp.stanford.edu/IR-book/html/htmledition/stemming-and-lemmatization-1.html.

14. Indurkhya, N. & Damerau, F. J. Handbook of Natural Language Processing, Second Edition. (Chapman and Hall/CRC, 2010).

15. Mikolov, T., Chen, K., Corrado, G. & Dean, J. Efficient Estimation of Word Representations in Vector Space. arXiv:1301.3781 [cs] (2013).

16. McInnes, L., Healy, J. & Melville, J. UMAP: Uniform Manifold Approximation and Projection for Dimension Reduction. arXiv:1802.03426 [cs, stat] (2020).

17. Ackermann, M. R., Blömer, J., Kuntze, D. & Sohler, C. Analysis of Agglomerative Clustering. Algorithmica 69, 184–215 (2014).

18. Breiman, L. Random Forests. Machine Learning 45, 5–32 (2001).

19. Fawcett, T. An introduction to ROC analysis. Pattern Recognition Letters 27, 861–874 (2006).

20. Lundberg, S. & Lee, S.-I. A Unified Approach to Interpreting Model Predictions. (2017).

21. Pedregosa, F. et al. Scikit-learn: Machine Learning in Python. Journal of Machine Learning Research 12, 2825−2830 (2011).

22. Candito, M. et al. Deep Syntax Annotation of the Sequoia French Treebank. In International Conference on Language Resources and Evaluation (LREC) (2014).

23. Jardim, D. L., Groves, E. S., Breitfeld, P. P. & Kurzrock, R. Factors associated with failure of oncology drugs in late-stage clinical development: A systematic review. Cancer Treat Rev 52, 12–21 (2017).

24. Malek, E. et al. Predicting Successful Phase Advancement and Regulatory Approval in Multiple Myeloma From Phase I Overall Response Rates. JCO Clin Cancer Inform 1, 1– 14 (2017).

25. Oxnard, G. R. et al. Response Rate as a Regulatory End Point in Single-Arm Studies of Advanced Solid Tumors. JAMA Oncol 2, 772–779 (2016).

26. Beinse, G. et al. Prediction of Drug Approval After Phase I Clinical Trials in Oncology: RESOLVED2. JCO Clinical Cancer Informatics 1–10 (2019) doi:10.1200/CCI.19.00023.

27. Adashek, J. J., LoRusso, P. M., Hong, D. S. & Kurzrock, R. Phase I trials as valid therapeutic options for patients with cancer. Nature Reviews. Clinical Oncology 16, 773– 778 (2019).

28. Christakis, N. A. & Lamont, E. B. Extent and determinants of error in doctors’ prognoses in terminally ill patients: prospective cohort study. BMJ 320, 469–472 (2000).

29. Vaswani, A. et al. Attention Is All You Need. arXiv:1706.03762 [cs] (2017).

30. Devlin, J., Chang, M.-W., Lee, K. & Toutanova, K. BERT: Pre-training of Deep Bidirectional Transformers for Language Understanding. arXiv:1810.04805 [cs] (2019).

